# Impact of seasonal malaria chemoprevention timing on clinical malaria incidence dynamics in the Kedougou region, Senegal

**DOI:** 10.1101/2024.04.16.24305915

**Authors:** Betty Kazanga, El-Hadj Ba, Eva Legendre, Mady Cissoko, Laurence Fleury, Lucas Bérard, Abdoulaye Diop, Cheikh Sokhna, Fode Danfakha, Issaka Sagara, Jean-Louis Ndiaye, Jean Gaudart, Jordi Landier

## Abstract

Seasonal malaria chemoprevention (SMC) with sulfadoxine-pyrimethamine and amodiaquine is recommended by the World Health Organization since 2012 for clinical malaria prevention in children in the Sahelian region of Africa. In Senegal, SMC implementation began in 2013 and is given to children under 10 years old. This study aimed to describe clinical malaria incidence in the general population during routine SMC implementation and to analyse how SMC timing impacted clinical malaria dynamics in eligible children.

We conducted an ecological study in the Kedougou region of Senegal in 27 villages included in the Bandafassi Health and Demographic Surveillance System (HDSS). We calculated weekly *Plasmodium falciparum* malaria incidence by age group using malaria case data recorded by community health workers and health-posts, and population denominators obtained from Bandafassi Health and Demographic Surveillance System. We used negative binomial generalized additive multilevel models to analyse the incidence of clinical episodes in children under 10 years during the expected SMC prophylactic period and at the end of the transmission period.

Malaria incidence was strongly seasonal with a high transmission period starting in June. Children under SMC presented an overall lower incidence compared to older children and young adults. Among children eligible for SMC, the incidence was lowest for approximately 3 weeks after treatment administration and increased subsequently, suggesting a gradual loss of protection. At the end of the high transmission period, a higher malaria incidence was recorded from the 3^rd^ to 6^th^ week after the week of administration of the fourth (final) SMC round.

While protecting children under 10 years, SMC warrants adjustment to reduce exposure before the next round, to increase protection of 5-9 years, and to cover the high transmission period completely. The addition of a 5^th^ round of SMC in 2023 was necessary to cover the end of the transmission season, but individual-level studies are required to ensure that drug efficacy and adequate dosing are maintained.

## Introduction

Seasonal malaria chemoprevention (SMC) has been recommended by the World Health Organization (WHO) since 2012 for the prevention of clinical malaria cases in children in the Sahel region of Africa where transmission is high and seasonal (1). It consists of a full treatment course of sulfadoxine-pyrimethamine (SP) and amodiaquine (AQ) given at monthly intervals during the high transmission season (2). Initially, SMC was recommended for children under the age of 5 years but has been extended to include children up to 10 years in Senegal (3). In Senegal, SMC has proven to be effective in reducing malaria incidence directly in targeted age groups and indirectly in non-target age groups by 60% and 26% respectively (4). Furthermore, SMC has been associated with a reduction of 45% in the incidence of severe malaria and of over 42% in-hospital deaths due to malaria (4,5).

Senegal is a malaria-endemic country located in the Sahel region of Africa with a population of 17,2 million inhabitants in 2021 (6). Malaria transmission is strongly seasonal with a high transmission season that coincides with the rainy season, from May to November, with regional variations (7). From 2015 to 2019, the country recorded a 38% reduction in malaria incidence and a 52% reduction in malaria-related deaths, in the context of increasing access to malaria diagnostic and treatment (8). However, progress appears to have stalled since 2020, with increasing incidence (26.7 cases/1000 in 2020 to 31.2 in 2021) and stable mortality (2.3 deaths per 100,000) (6). This increase was attributed to the impact of the COVID-19 pandemic and longer rains (9). The burden of malaria is now largely concentrated in the 3 southeastern regions of Kedougou, Kolda, and Tambacounda, with respective incidence rates of 537, 150 and 215 cases/1000 in 2021. All other regions reported incidence <10 cases/1000 (6).

In Senegal, the implementation of SMC began in 2013 and is led by the National Malaria Control Program (NMCP). Community health workers (CHW) administer monthly SMC to children between 3 months to 10 years of age during the high transmission season. All 3 days of treatment are directly observed by CHW with a door-to-door approach (12). SMC is currently conducted in 16 high-burden districts of 5 regions (12). The number of rounds in the campaign is defined by region and depends on the duration of the high transmission season. In 2020 and 2021, there were 4 rounds administered in the Kedougou region and 3 rounds in the other selected areas with a campaign coverage of 95% and 87% respectively (9,10). In addition to SMC, the current malaria control strategies in Senegal are free early diagnosis and treatment available through community health workers and health posts, vector control (long-lasting insecticide-treated nets and indoor residual spraying in 2020 and 2021), and intermittent preventive treatment in pregnancy (IPTp) (9).

Despite the deployment at high coverage of all these recommended malaria strategies in Senegal, the country has not yet attained zero-malaria status. Therefore, optimizing ongoing interventions to reduce the burden of malaria in high-incidence regions is a critical step toward country-level elimination. Previous studies conducted in Sahelian countries have shown that SMC protects children for 28 days and thus requires a strict timing of SMC programs with repeated treatments at monthly intervals to achieve optimal results (2,11). Other studies suggested that the number of rounds should be adapted to the duration of the high transmission season(12) Indeed, the Senegalese NMCP reported that >50% of malaria cases in 2020 occurred after the final round of SMC despite high coverage of SMC (9,13), suggesting that the 4 rounds may not provide adequate cover throughout the entire high-risk season. In addition, resistance to sulfadoxine-pyrimethamine remains a major threat to SMC effectiveness (14). Therefore, this study evaluated the dynamics of clinical malaria incidence in relationship with the timing of SMC drug delivery in the Kedougou region of Senegal during the 2020 and 2021 SMC campaigns. The aim was to determine how it can be fine-tuned to maximize the impact of the intervention in terms of malaria incidence reduction.

## Methods

### Study design and setting

This ecological study of *P. falciparum* clinical malaria incidence was conducted in the Kedougou region, bordering Mali and Guinea, 700km from Dakar. The regional population was estimated around 200,000 inhabitants in 2021 (3,6). We included villages covered by the Bandafassi Health and Demographic Surveillance System (HDSS) and within the catchment area of two health posts, Thiabedji and Bandafassi (15). The main ethnic groups in the study villages are Bedik, Mandinka, and Fula Bande. Access to malaria diagnosis and treatment is available from Thiabedji and Bandafassi health posts, as well as community health workers (DSDOM (dispensateurs de soins à domicile, i.e. "home-based care providers") and ASC (agents de santé communautaire, i.e. "community health workers") (10) in most villages. In this study, we use community health workers (CHW) to refer to DSDOM and ASC because they provide similar malaria diagnosis and treatment at the community level in Senegal using rapid diagnostic tests (RDT) and treating uncomplicated malaria with artemisinin-based combination therapy (ACT) while referring severe malaria, and malaria cases in children under 5 or pregnant women to the nearest health post.

### Malaria case data

At the beginning of each year, RDT-confirmed *P. falciparum* malaria individual case data was collected from consultation registries of CHW and health posts for the previous year and entered in Microsoft Excel. The individual patient information was anonymized and included age, sex, address, date of consultation, RDT result and treatment provided.

### Population data

Population data by age group and village used in this study was collected by the Bandafassi Health and Demographic Survey System (HDSS) in 2020 and 2021 (15). Since 2019, the population data is updated through yearly home visits by field survey teams, who record the status of all household members by interviewing an adult of the household who was present all year long to record births, deaths, newly arrived members, or those who left.

### Ethics

This study is included in the "Malaria Asymptomatic Reservoir in the Sahel" research project and its protocol was approved by the National Ethics Committee for Health Research of Senegal (protocol SEN20/10, N°0000052/MSAS/DPRS/CNERS)

Since 2020, a written formal consent was obtained from each head of household or his/her representatives by signing an informed consent form allowing the collection of demographic data by the HDSS. The written formal consent is valid for several rounds. In 2021 and 2023, new consent forms are signed in cases where there is a new household or when the head of the household has changed.

### SMC campaign

In 2020 and 2021, the SMC campaigns consisted of 4 rounds from the end of June to early October (Table 1)

**Table 1.**
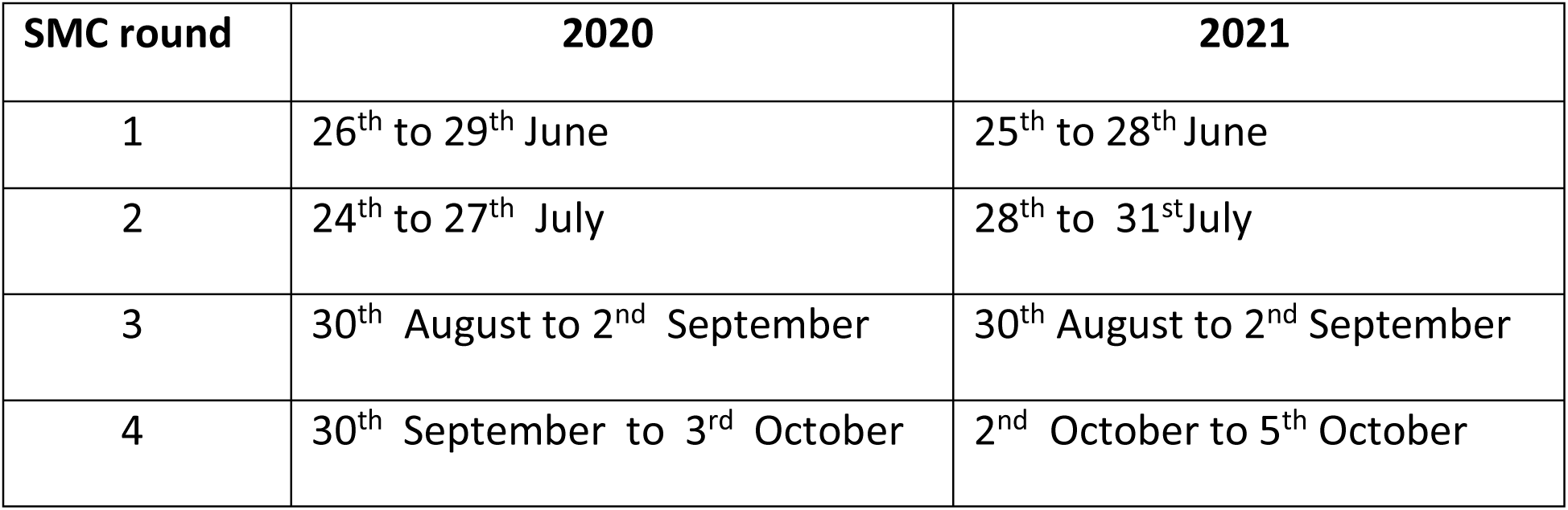
Dates of beginning and end of SMC rounds in 2020 and 2021.

| SMC round | 2020 | 2021 |
| --- | --- | --- |
| 1 | 26<sup>th</sup> to 29<sup>th</sup> June | 25<sup>th</sup> to 28<sup>th</sup> June |
| 2 | 24<sup>th</sup> to 27<sup>th</sup> July | 28<sup>th</sup> to 31<sup>st</sup> July |
| 3 | 30<sup>th</sup> August to 2<sup>nd</sup> September | 30<sup>th</sup> August to 2<sup>nd</sup> September |
| 4 | 30<sup>th</sup> September to 3<sup>rd</sup> October | 2<sup>nd</sup> October to 5<sup>th</sup> October |

### Study population selection

#### Inclusion criteria

This study included all individuals matching the following criteria: *P.falciparum* RDT positive (malaria cases), date of diagnosis documented in 2020 or 2021, age documented, and residing in a village included in the Bandafassi HDSS and within the catchment area of Bandafassi and Thiabedji health posts.

#### Exclusion criteria

Individuals from unknown and out-of-zone villages were excluded. In addition, individuals from villages within the study area that had incomplete malaria case data (eg missing age) were also excluded. Villages within the Bandafassi and Thiabedji health post catchment area that had no functional CHW serving the village or no documentation of malaria cases by health workers for an extended period (up to months) in a year were also excluded after identification due to low malaria cases counts and confirmation of a local health personnel

### Data analysis and statistical methods

All analysis was conducted using R software version 4.2.2, and {mgcv} package for the GAMM analysis.

### Descriptive analysis

#### Malaria incidence

Malaria incidence was expressed as the number of cases per 1000 population per week. Based on this descriptive analysis and a previous study assessing climate-driven variations in malaria transmission in Senegal (16), the years were divided into 2 transmission periods: low transmission period and high transmission period. Incidence was calculated for 5-year age groups for individuals <20 years, using age-group and village**-**specific denominators, including both 0-4 and 5-9 eligible for SMC and 10-14 and 15-19 uneligible for SMC.

### Statistical analysis

#### Variable definition

The weeks after the SMC round variable is defined as the number of weeks that passed after the administration of the SMC round. Week 0 therefore corresponds to the week in which SMC was administered and week 1, is the first week after the week of the SMC round and so forth (S1 Fig). We defined the week of the year as the calendar week (1–52), and this variable was used to adjust for varying incidence levels across a given season (Fig 5).

#### Evaluation of SMC protection during the expected prophylactic period

This analysis was restricted to children targeted with SMC i.e. 3 months to <10 years (or 3-120 months) and to the period between the first day of the first round of SMC to 5 weeks after the 4^th^ and final round. The outcome variable was the weekly count of clinical malaria cases diagnosed per age group and village over time. Explanatory variables were the number and weeks elapsed since the latest SMC round, age group, week of the year, and the year (2020 or 2021). A negative binomial generalized additive multilevel model including the log-transformed person-weeks which was added as an offset was used to estimate incidence rate ratios (IRR) and a 95% confidence interval (CI). Generalized Additive models were used to rely on smoothing functions to adjust for temporal (week of the year, univariate spline) autocorrelations, without assuming a specific shape of the relationship. Random intercepts were included to account for multiple non-independent observations for each village (indexed by i) and for each age group within a village (indexed by j). The default thin plate regression spline settings were used for the smooth term (f).

The final model for this analysis is given in Equation 1 as follows:

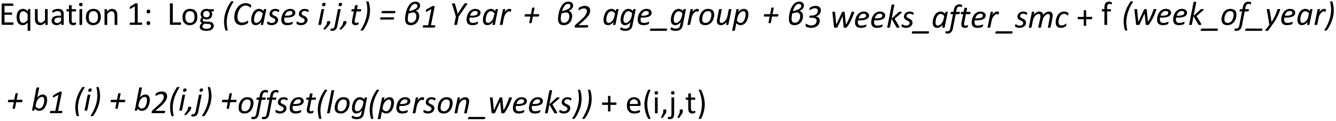

*Cases i,j,t* represent the number of clinical malaria cases diagnosed per village *(i)* and age group *(j)* over time (*t*), β’ s are the parametric coefficients, f is the smooth (spline) functions, while b’s are the random effects, log(person-weeks) is the offset and e is the error term

#### Evaluation of SMC protection at the end of the transmission period

This analysis was also restricted to malaria cases and the population of children targeted with SMC i.e. 3 months to <10 years (or 3-120 months) and to the period following the final SMC round, from week 0 of the 4^th^ SMC round to the end of the year. The aim was to evaluate if the 4-round SMC campaign extended effective protection until transmission reached sufficiently low levels. Univariate and multivariate GAMM similar to the previous analysis were used. The outcome variable was the weekly count of clinical malaria cases diagnosed per age group and village over time. The explanatory variables were the year of case diagnosis, age group of cases, and weeks after SMC round 4. Random intercepts were included to account for multiple non-independent observations for each village and each age group within a village. The log-transformed person weeks were added as an offset. In addition, a negative binomial model was used to estimate incidence rate ratios (IRR) and a 95% confidence interval (CI).

The final model for this analysis is given in Equation 2 as follows:

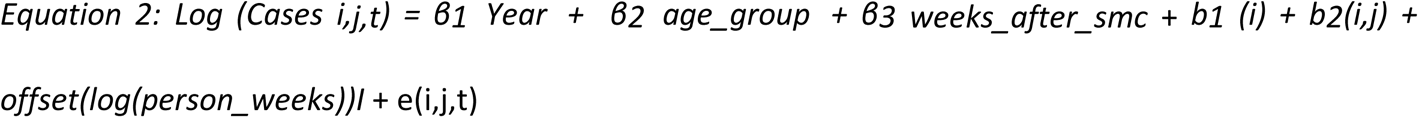

## Results

### Data selection

After data entry, we obtained 8,480 confirmed malaria cases diagnosed by 29 CHWs and 2 health posts in 31 villages within the study area in 2020 and 2021 (Fig 1). After applying exclusion criteria, 3 426 clinical cases in 25 villages with a population of 8910 in 2020 and 3 319 clinical cases from 27 villages with a population of 9 348 were added in this study. Overall, 15.7% (n=1062) of cases and approximately 25% of the population were under 10 years of age.

**Fig 1.**
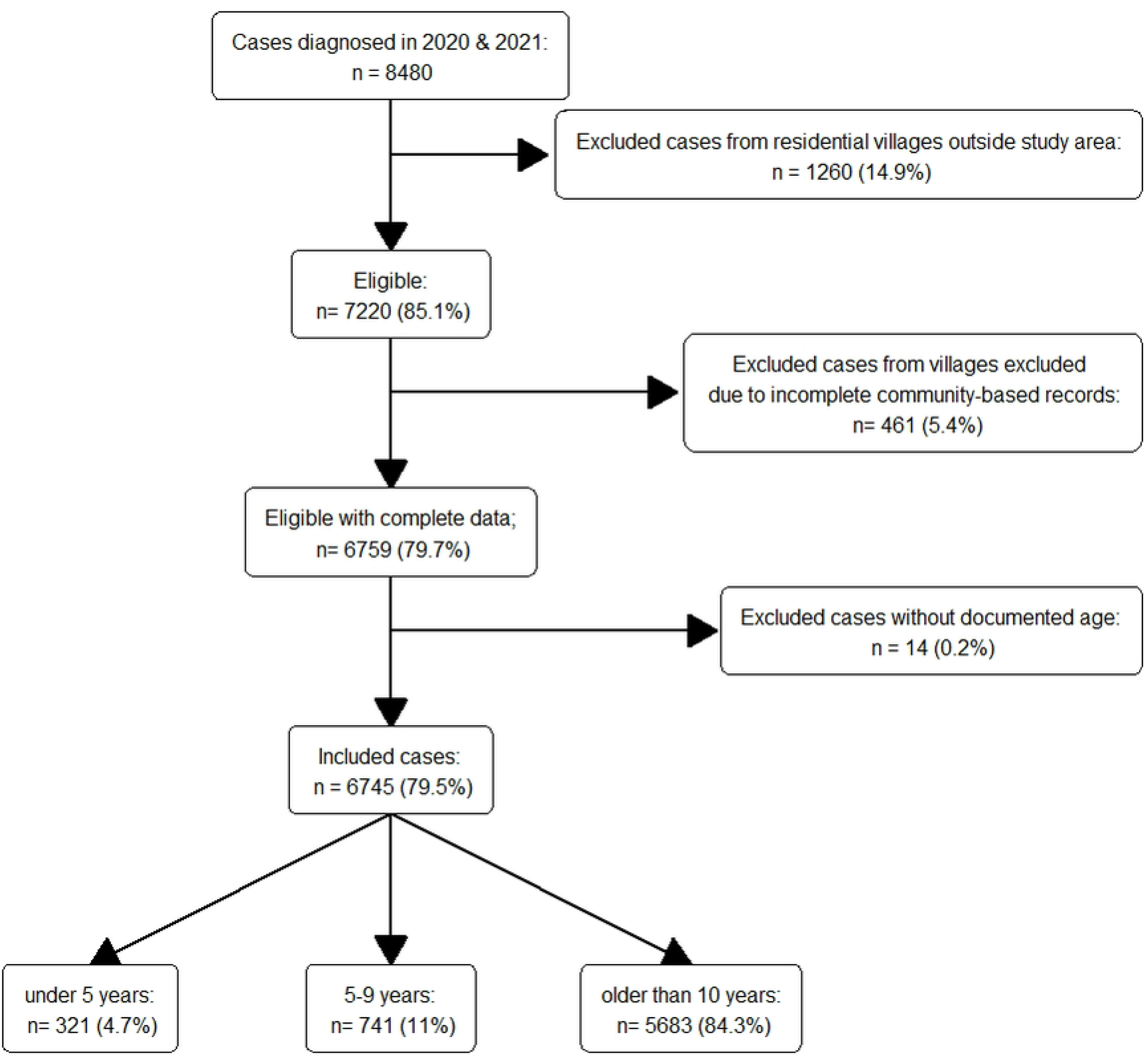
Flow chart of data selection. In 2020, 6 villages were excluded due to incomplete community-based records and 4 villages in 2021 (461 cases in total).

### Incidence of malaria

The annual malaria incidence in the study area was 385 and 355 cases per 1000 population in 2020 and 2021 respectively. The time series of malaria incidence showed that incidence was seasonal in both years, with the high transmission beginning in June, and peak incidence reaching early August for both years (Fig 2). In 2021, malaria incidence persisted longer than in 2020, with a slower decrease and persistence until the end of December (Fig 2).

**Fig 2:**
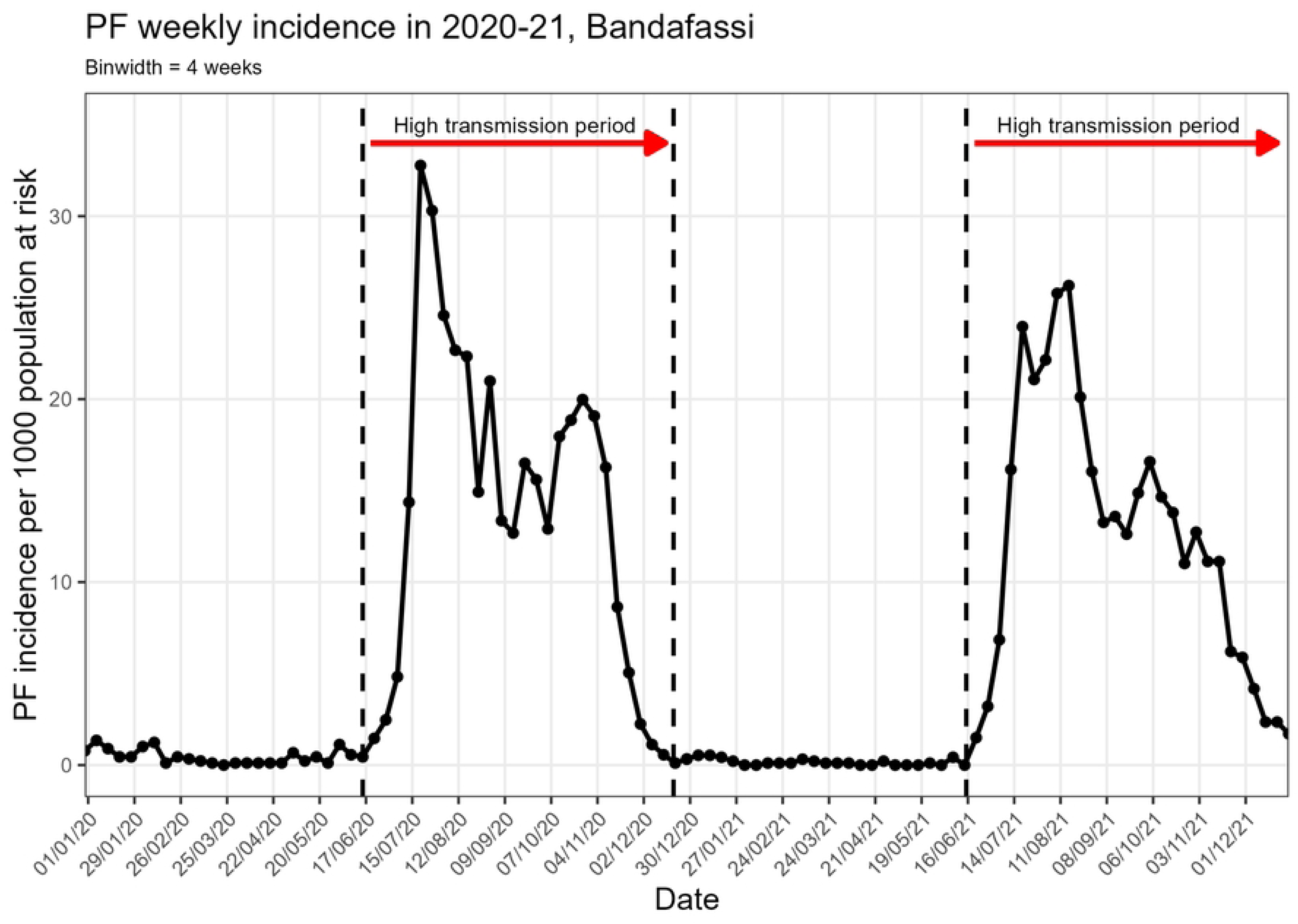
Weekly malaria incidence per 1000 population at risk in Bandafassi HDSS area, 2020-21.

Children aged 0-4 (covered by SMC) had the lowest incidence, while individuals aged 10-14 and 15-19 had the highest (Fig 3, Fig S2).

**Fig 3:**
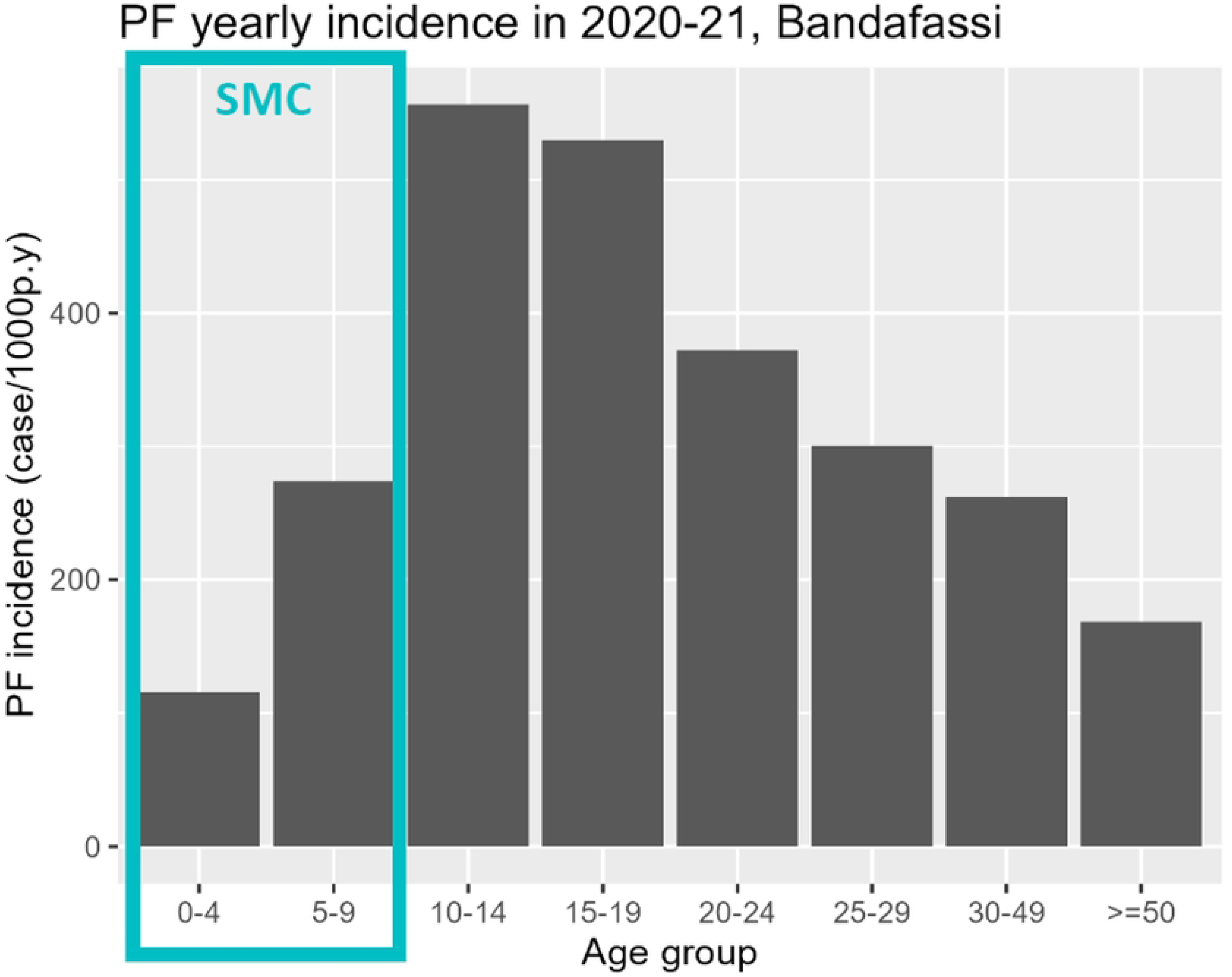
Yearly malaria incidence per 1000 population at risk in Bandafassi HDSS area, 2020-21.

### Timing and duration of protection provided for each SMC round

The interval between the SMC rounds in 2020 was not constant and ranged from 27 to 36 days. In contrast, the interval between the rounds in 2021 was constant with 32 days between each round (S1 Fig). In children 0-4 and 5-9, incidence appeared to fluctuate between SMC rounds with lower incidence in the weeks immediately after the SMC distribution, and an increase 1-2 weeks before the next round. (Fig 4) This pattern was specific to SMC eligible groups, and not identified among 10-14 or 15-19 years old.

**Fig 4:**
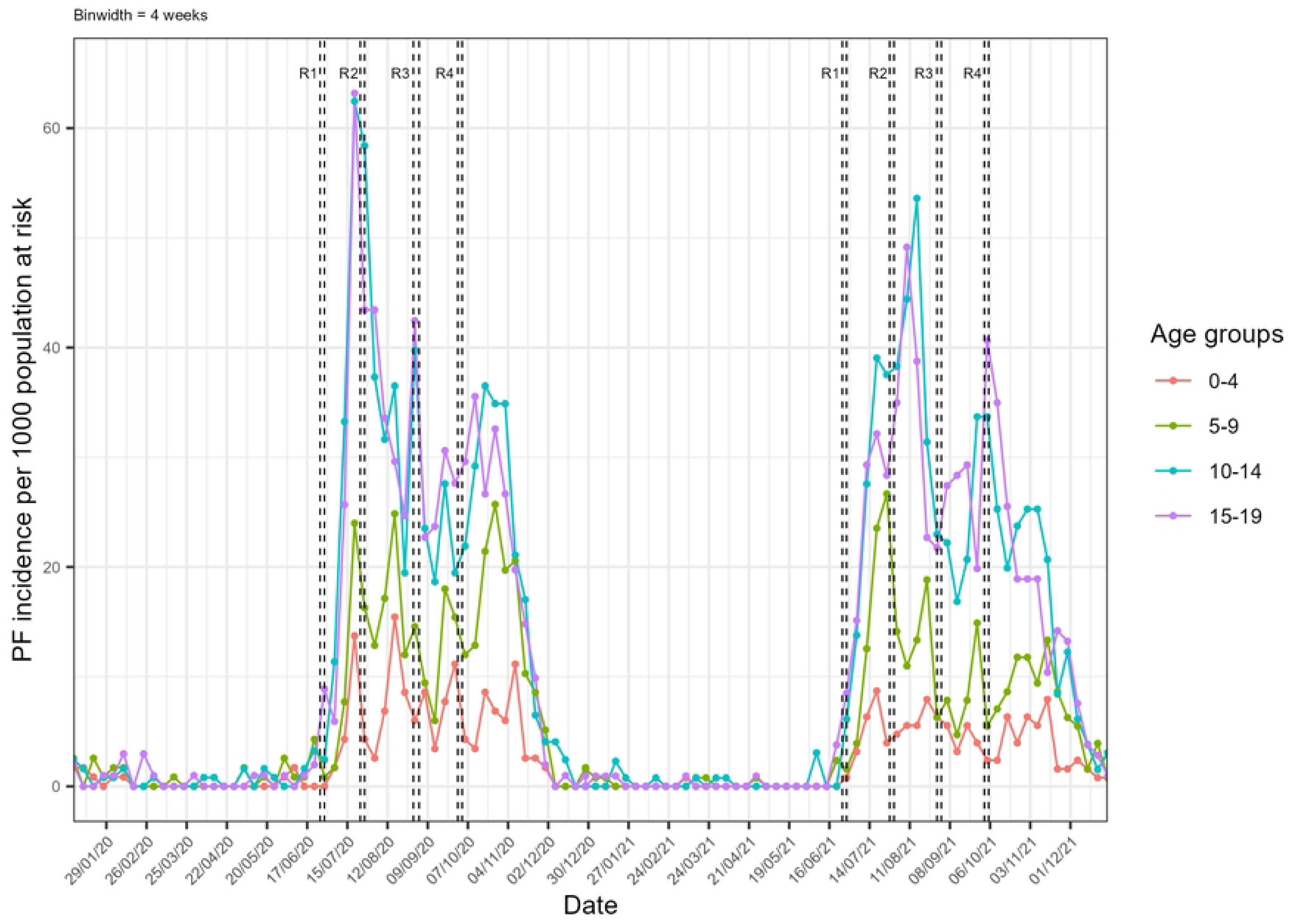
Weekly incidence by age groups (<20 years) in the Bandafassi HDSS area (2020-2021). The SMC round number is represented by the letter R followed by the round number. The vertical dashed lines represent the start and end dates for SMC treatment during each round.

The start of the high transmission period was not uniform across all age groups in both years with some of the age groups having a slower start (e.g. 0-4 years) (S2 Fig). Likewise, all groups presented an increase towards the end of the high transmission season, but the timing differed between SMC-eligible and ineligible groups.

### SMC protection during the expected prophylactic period

After adjustment on year, age group, and week of the year, there was no statistically significant difference in risk from week 0 (week starting on the first day of the SMC round) to week 2, i.e. during the first 3 weeks since the start of each SMC round (Fig 5, S1 Table). The risk increased thereafter from week 3 and was highest during week 5 after SMC (IRR 2.77, 95% CI 1.62-4.72). In this analysis, malaria risk was lower in 2021 (adjusted IRR= 0.70, 95% CI 0.60-0.83) compared to 2020, and children aged 5-9 years had a statistically significant higher risk of malaria (IRR= 2.15, 95% CI 1.75-2.65) compared to children under 5 (S1 Table). The univariate spline for the week of the year exhibited oscillations between SMC rounds and corresponding to increasing risk occurring before the deployment of the next round from approximately 3 weeks onwards(Fig 6a). In contrast, after adjustment of the week since SMC, the multivariate spline matched the shape of the general malaria incidence (Fig 6b, Fig 2), suggesting good adjustment over seasonal transmission dynamics. A model using the SMC period variable was also tested and gave the same results for increasing risk from week 3 onwards.

**Fig 5:**
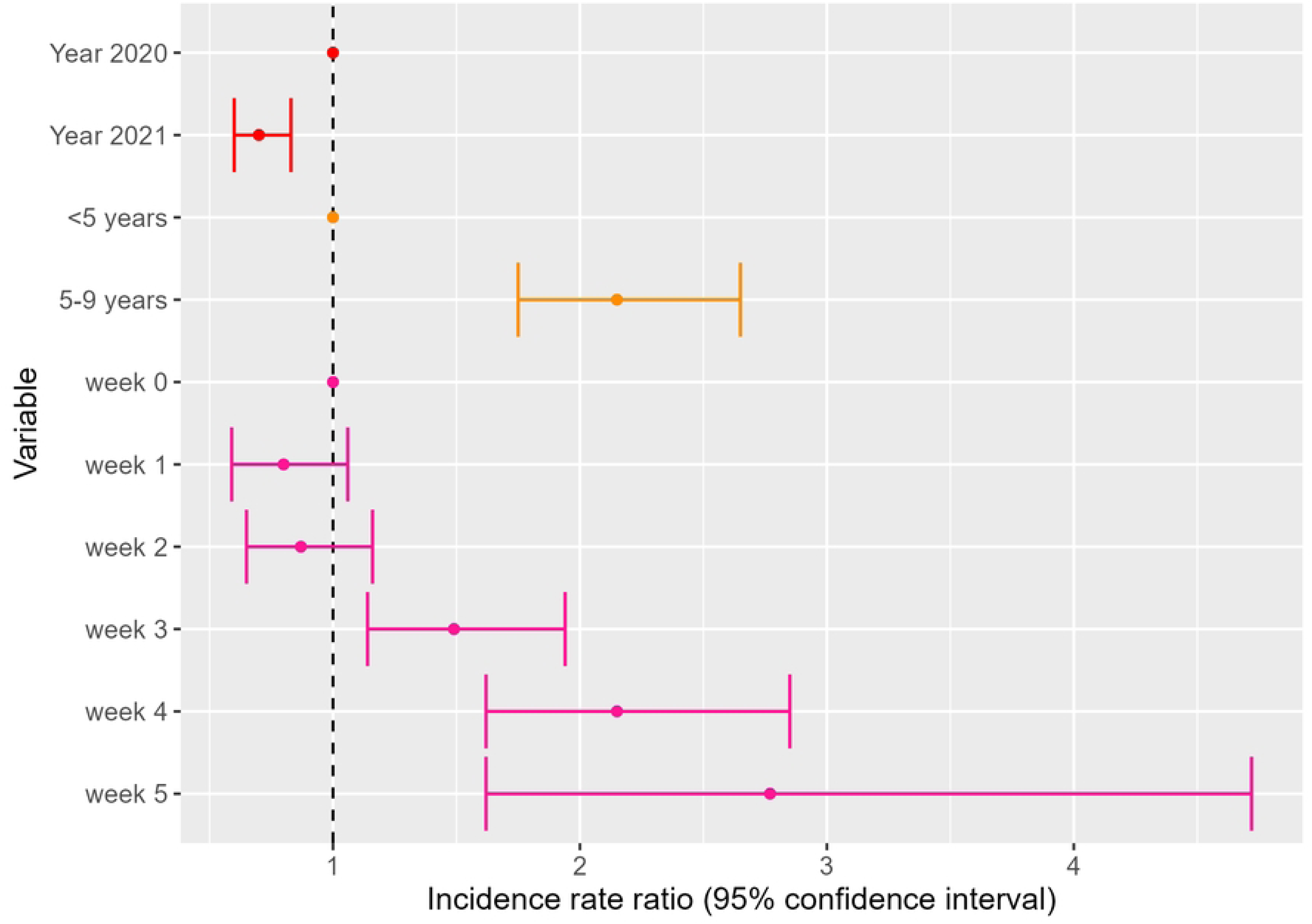
Results of the multivariate GAMM evaluating the relationship between malaria incidence and year of diagnosis, age groups receiving SMC, and weeks after SMC round.

**Fig 6:**
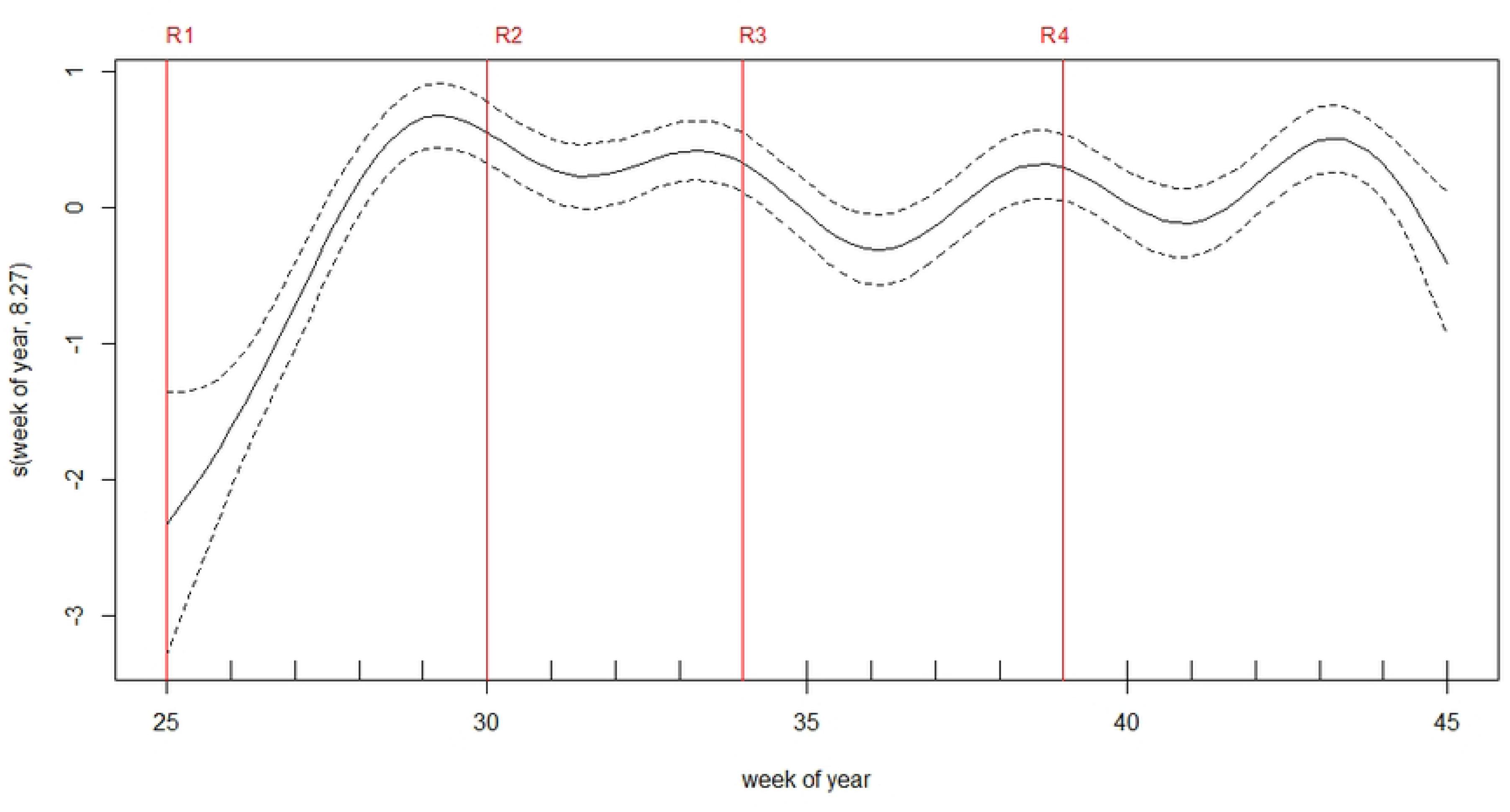
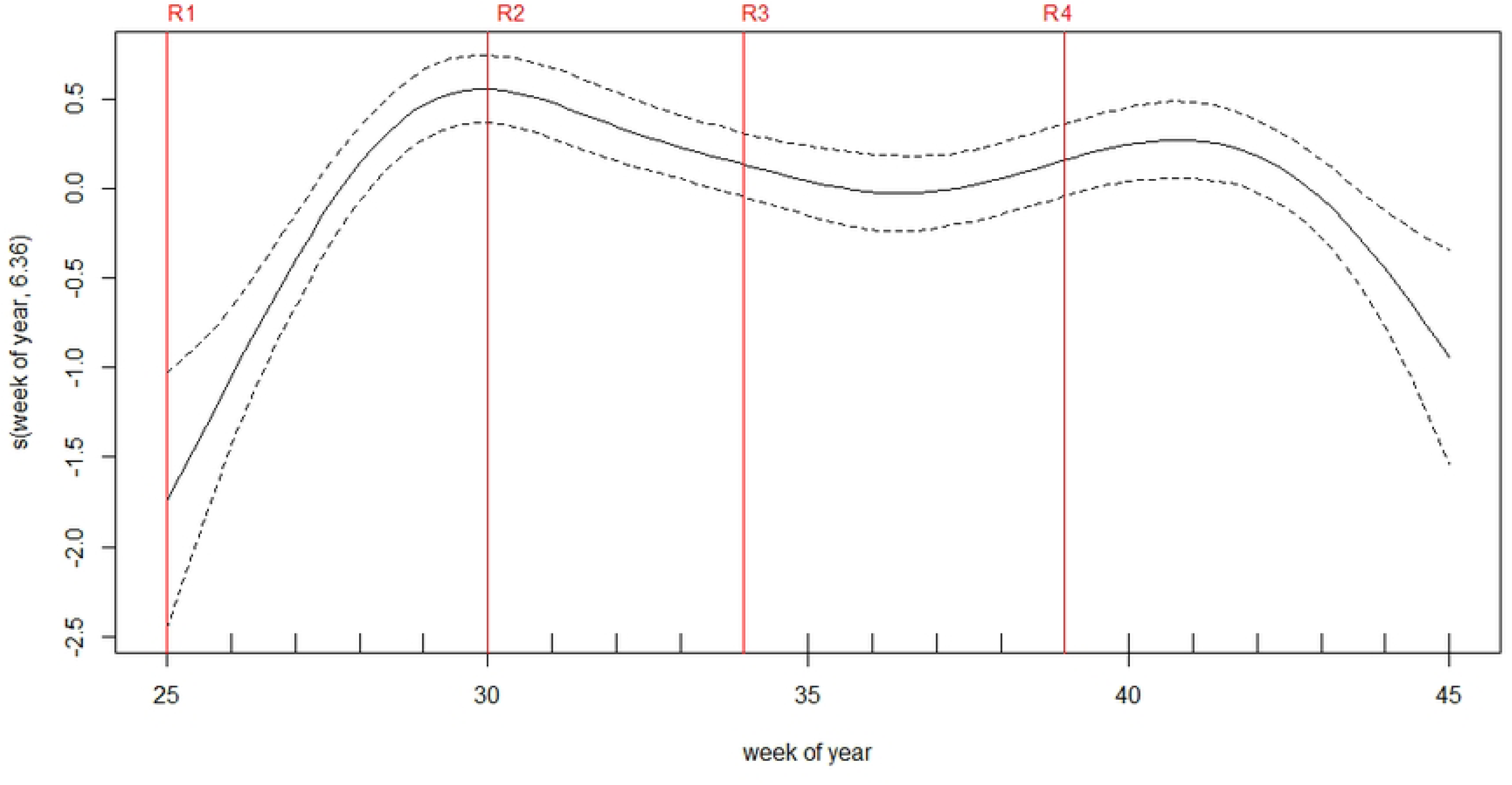
Smoothing components of week number in the GAMM. The week of the year an SMC round was administered is represented by a red vertical line (R1: round 1…). This "week of year" variable provides a temporal adjustment. (a) Spline for the week of the year in the univariate model, displaying incidence oscillations. (b) Spline for the week of the year in the multivariate model: after adjusting for the number of weeks since SMC, the spline presents with the overall shape of the general population incidence as presented in Fig 2. Week 25 corresponds to the week of 22^nd^ and 21^st^ June 2020 and 2021 respectively which is the onset of the SMC campaign.

**Fig 7:**
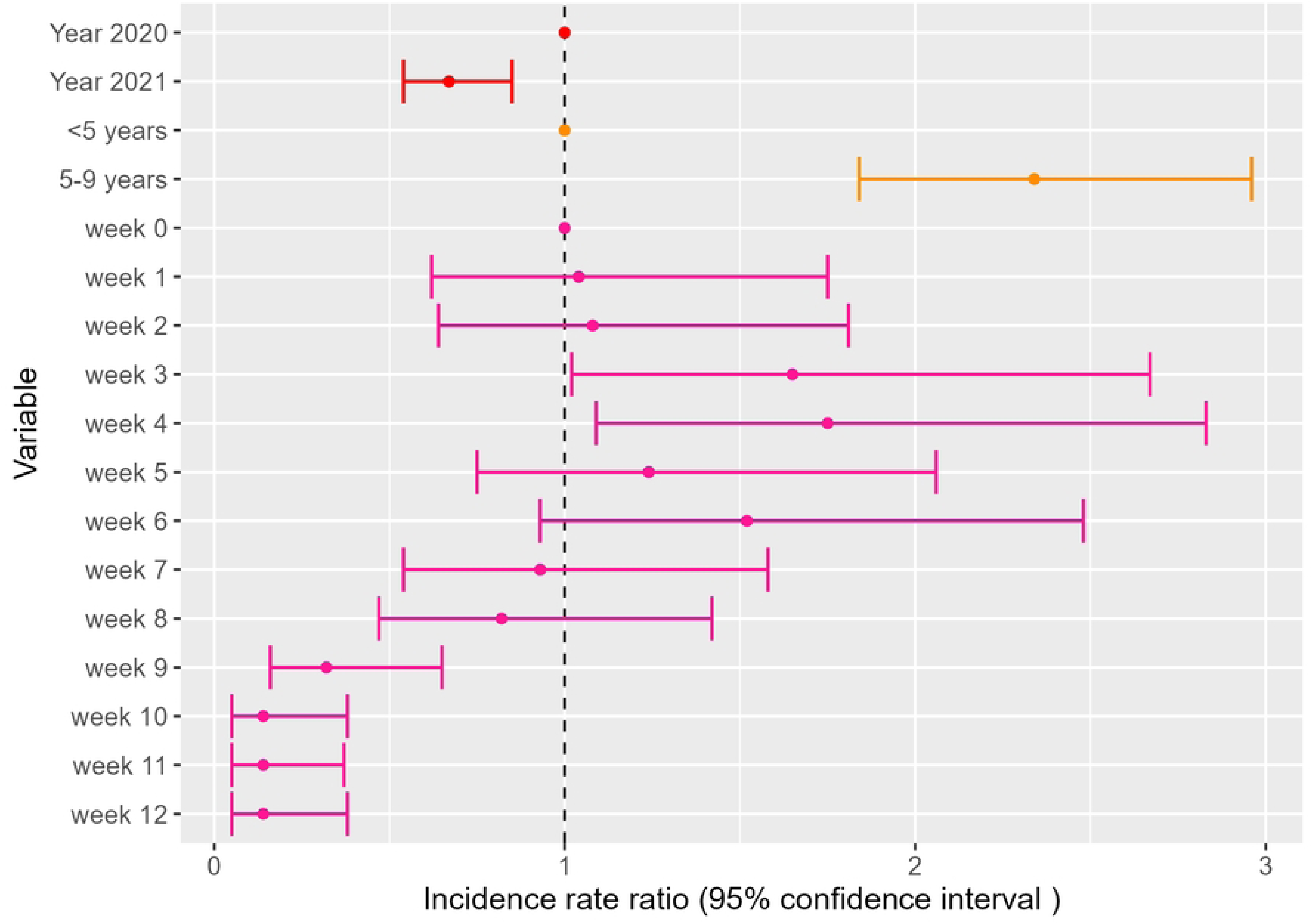
**Multivariate GAMM evaluating the relationship between malaria incidence and year of diagnosis, age groups receiving SMC and week after SMC.**

### SMC protection at the end of the transmission period

After adjusting for year, age group, and number of weeks after SMC, there was no statistically significant difference in risk from week 0 to week 2 after SMC round 4. Malaria risk was persistently higher from week 3 till week 6 after SMC with the highest risk during week 4 (IRR 1.75 95% CI 1.09-2.83). Thereafter, there was a decrease in risk from week 7 to the end of the year where the risk was lowest (week 10-12) with an IRR of 0.14 (95% CI 0.05-0.38). Similarly to the previous model, the 2021 risk was lower and children aged 5-9 had higher risk (Fig7, S2 table).

## Discussion

In this study, malaria incidence in the Kedougou region of Senegal remains highly seasonal and shows large variations across age groups, in particular among younger individuals. While providing a significant reduction in incidence to eligible children, SMC protection was inconsistent for targeted age groups. At population level, protection started waning 3 weeks after treatment administration, which proved further detrimental when SMC rounds were inconsistently timed: the lowest risk was obtained from day 0 to day 20, a 50% increase from day 21 to day 27, a 100% increase from day 28 to day 34, and a 177% increase beyond. Furthermore, the round 4 protection waned too early to cover the end of the high transmission period. Lastly, children aged 5-9 had higher incidence levels compared to children 0-4.

This ecological study shows that, at the population group level, SMC protection wanes off 1 to 2 weeks before the next round An increased incidence was already reported when comparing the effect of SMC in children before and after 28 days since treatment uptake (2). Our study suggests that this is also taking place in the Senegalese context, where SMC is directly observed for all 3 days of treatment. Therefore, the longer intervals between rounds in 2020 likely contributed to the higher risk of malaria compared to 2021. The results also show that, in Kedougou, 4 rounds were insufficient to cover adequately the end of the transmission season. Thus, they fully support the decision of Senegalese NMCP to add a fifth round of SMC in the Kedougou region (9). Evidence-based improvement of SMC intervention allocation is also ongoing in several countries, to adapt the start and duration of SMC campaigns to regional malaria transmission patterns (12).

The lower protection observed for children aged 5-9 compared to younger children is unexpected: in an earlier clinical trial conducted in the nearby Saraya district in the Kedougou region, both groups experienced comparable incidence levels (17). The difference could originate from different malaria preventive behaviors, leading to lower exposure in younger children: children under 5 could enjoy better protection from long-lasting insecticide-treated nets (LLINs) than those aged 5-9 years since they usually sleep earlier and are closely monitored by their parents, while children aged 5-9 years are more likely to be outdoors in the evening. Alternatively, lower uptake of SMC among older children could also contribute to the higher baseline levels observed among 5-9-year-olds. We lack disaggregated SMC participation data to compare coverage of intervention precisely across age groups and villages, but health-area-level coverage reported by Bandafassi and Thiabedji health posts is high (>90%) and homogenous across all reported age groups (3-11 months, 12-59 months, 60-120 months) (13). If higher incidence among the 5-9 resulted was solely from lower coverage, it would not follow patterns matching the timing of SMC rounds, but rather seasonal patterns as observed in older individuals.

Beyond the difference between younger and older SMC-eligible children, the duration of protection from SMC appeared short, around 3 weeks. Individuals may be reaching drug levels permissive for parasite multiplication earlier than the expected four weeks, either due to increasing resistance levels in parasites or to insufficient dosing of drugs (18). In Senegal, SMC is administered under 3-day supervision, which increases drastically the number of children receiving the complete SP-AQ regimen including all 3 doses of AQ. Recent analyses on genetic markers of Plasmodium falciparum drug resistance reported evidence of selection on genetic loci associated with SP resistance(19).

Our results also highlight a high incidence observed in individuals aged 10-19 years old, who now represent the most affected group. We lack comparable retrospective data to understand if their higher burden results from a specific increase in incidence, or a lesser decrease compared to younger individuals, now targeted with SMC. Their exposure to SMC was very different: while the youngest were likely exposed to SMC throughout most of their childhood, the oldest may only have received it for a few years. The comparable incidence between the age groups suggests that multiple factors are at play. Children 10-19 are likely to have lower immunity compared to older individuals. They also may have higher exposure, likely due to different behavioral patterns such as nighttime outdoor activities, use and adherence to preventive interventions (e.g. SMC, insecticide-treated nets), and mobility patterns that make an individual susceptible to clinical infection.

The key strength of this ecological study of clinical malaria incidence was that it relied on a wealth of data available through a combination of routinely collected malaria case data and high-precision population denominators at the age-group level obtained from the yearly census conducted in the Bandafassi DHSS. The routine individual case data combined with yearly updated census data allowed studying weekly incidence by age group and village. Such analyses are usually limited by the availability of up-to-date census in these settings. In addition, the context of community-based, free access to diagnostic and treatment alleviates geographic and financial barriers to seeking care, which provides a less biased evaluation of the clinical burden in the general population (20). This study also demonstrates how disaggregated routine clinical malaria data or aggregated by 5-year age groups enables the identification of relevant signals such as age-specific incidence dynamics and consistency of SMC protection. Our analysis assumed a constant denominator during a given transmission season, which suggests that analyzing the dynamics of well-documented case time series (e.g. from sentinel health facilities) alone could provide relevant insights to monitor changes in SMC effectiveness. The main limitation is the absence of village-level SMC coverage data, which could not be retrieved retrospectively as done for consultation registries. While health-area level coverage appears homogenous across age groups and rounds, village and round-specific participation data could have improved our assessment of SMC-associated incidence dynamics. Furthermore, in this study, the week of the year was used to adjust for seasonal changes in malaria risk. In other studies, these seasonal changes can be modeled using meteorological parameters, such as rainfall, temperature, and/or composite indicators, and their effect on incidence can be quantified (16,21).

Beyond the description of ranges of improvement of SMC, this study highlights the persistence of *P. falciparum* transmission in the Kedougou region despite a high level of coverage with control measures. While deploying SMC with SP-AQ every 3 weeks (before the protection wanes off) may not be feasible, efforts should be made to maintain the 30-day interval between rounds as a maximum to reduce exposure before the next round. Our results also highlight the need to assess SP-AQ efficacy to provide definitive evidence regarding the need for an alternative treatment, more efficient than SP-AQ and lasts for 4 weeks or longer (18). In addition, other malaria control strategies such as seasonal vaccination in younger children may be considered to be given alongside SMC and provide cumulative protection (22). However, it is critical to consider the cost-effectiveness of deploying additional control strategies to children <10 years considering that the largest burden of cases now affects mainly older age groups, despite vector control measures though limited. The results of the 2020-2021 malaria indicators survey in Senegal show that only 66.5% of the population in the Kedougou region had access to an insecticide-treated net in the household (23). Indoor residual spraying was undertaken in Kedougou in 2020 and 2021 but discontinued in 2022. Designing effective additional vector control interventions requires an evaluation of outdoors and residual transmission: outdoor biting was reported at high levels in 2019 (24). After nearly two decades of indoor vector control, it is likely to have further increased.

Individuals aged 10 years and older now support the majority of the clinical malaria burden, but they are also likely responsible for *P. falciparum* persistence and circulation across dry and wet seasons. In addition, there are new risk groups that have been identified such as gold miners, transhumants, and talibés in Qur’anic schools that can also benefit from targeted interventions. Designing and implementing interventions targeting individuals at high risk of parasite carriage could represent a relevant strategy to further decrease transmission in Kedougou and similar settings with a high degree of interventions already targeting groups at high risk of severe disease.

## Conclusion

Kedougou region now harbors the highest malaria incidence in Senegal despite significant epidemiological changes resulting from control activities over the past 15 years. SMC presents a striking effect on eligible children under 10 years, who no longer bear the heaviest clinical burden. While protecting children under <10 years, SMC timing could be improved to avoid increased incidence 1-2 weeks before the next round. Surveillance of incidence patterns among eligible groups in routine data could also contribute to monitoring SMC performance. Finally, malaria elimination strategies in this area should not overlook the shift in burden to older age groups, asymptomatic carriage, mobile individuals, and newly identified risk groups.

## Data Availability

We are currently in the process of uploading the data respectively required to produce figures 1-4 and figure 5-7 and corresponding models on the Senegalese Statistical agency repository.

https://www.ansd.sn/

## Acknowledgments

We would like to thank the population of Bandafassi, the CHW, and the health personnel of the Kedougou district and Bandafassi and Thiabedji health posts. We also thank Moussa Ba and Gérald Keita for their major help for onsite data collection, and Victorine for data entry at IRD Dakar. We acknowledge the interviewers and supervisors of Bandafassi HDSS census teams who collected the door-to-door, yearly updated population information.

For the purpose of Open Access, the authors have applied a CC BY public copyright license to any Author Accepted Manuscript version arising from this submission.

## Supporting information

**S1 Fig: Graphical definition of variables (not showing exact rounds or dates).**

The SMC round number is represented by the letter R followed by the round number. Each round has 3 days of directly observed treatment. Week 0 is the week in which SMC was administered. The "SMC period" variable was defined as the interval between SMC rounds. For instance, SMC period 1 corresponds to the time between round 1 and round 2. Between two rounds, the maximum duration of a period was 36 days. The duration of SMC period 4, corresponding to the last round of SMC was therefore set up to week 5.

**S2 Fig: Weekly incidence by age groups in Bandafassi HDSS area** (**2020–2021**).

The scale of the y-axis changes differs across panels.

**S1 Table: Multivariate multilevel GAM evaluating the relationship between malaria incidence and year of diagnosis, age groups receiving SMC, and week after SMC, during the expected prophylactic period after each SMC round.**

**S2 Table: Multivariate multilevel GAM evaluating the relationship between malaria incidence and year of diagnosis, age groups receiving SMC, and week after SMC, during the period starting with the 4^th^ and final SMC round and finishing at the end of the year.**

**S1 Checklist: STROBE checklist**

